# Clotting factor genes are associated with preeclampsia in high altitude pregnant women in the Peruvian Andes

**DOI:** 10.1101/2021.05.20.21257549

**Authors:** Keyla M. Badillo Rivera, Maria A. Nieves-Colón, Karla Sandoval Mendoza, Vanessa Villanueva Dávalos, Luis E. Enriquez Lencinas, Jessica W. Chen, Elisa T. Zhang, Alexandra Sockell, Patricia Ortiz Tello, Gloria Malena Hurtado, Ramiro Condori Salas, Ricardo Cebrecos, José C. Manzaneda Choque, Franz P. Manzaneda Choque, Germán P. Yábar Pilco, Erin Rawls, Celeste Eng, Scott Huntsman, Esteban González Burchard, Giovanni Poletti, Carla Gallo, Carlos D. Bustamante, Julie C. Baker, Christopher R. Gignoux, Genevieve L. Wojcik, Andrés Moreno-Estrada

## Abstract

**Study question:** What is the genetic basis of preeclampsia in Andean families residing at high altitudes?

**Summary answer:** A top candidate region associated with preeclampsia containing clotting factor genes *PROZ, F7* and *F10* was found on chromosome 13 of the fetal genome in affected Andean families.

**What is known already:** Preeclampsia, a multi-organ complication of pregnancy, is a leading cause of maternal morbidity and mortality worldwide. Diagnosed by the onset of maternal hypertension and proteinuria after 20 weeks of gestation, this disorder is a common cause of preterm delivery and affects approximately 5-7% of global pregnancies. The heterogeneity of preeclampsia has posed a challenge in understanding its etiology and molecular basis. However, risk for the condition is known to increase in high altitude regions such as the Peruvian Andes.

**Study design, size, duration:** To investigate the genetic basis of preeclampsia in a high-altitude resident population, we characterized genetic diversity in a cohort of Andean families (N=883) from Puno, Peru, a high-altitude city above 3,500 meters. Our study collected DNA samples and medical records from case-control trios and duos between 2011-2016, thus allowing for measurement of maternal, paternal, and fetal genetic factors influencing preeclampsia risk.

**Participants/materials, setting, methods:** We generated high-density genotype data for 439,314 positions across the genome, determined ancestry patterns and mapped associations between genetic variants and preeclampsia phenotype. We also conducted fine mapping of potential causal variants in a subset of family participants and tested ProZ protein levels in post-partum maternal and cord blood plasma by ELISA.

**Main results and the role of chance:** A transmission disequilibrium test (TDT) revealed variants near genes of biological importance in pregnancy physiology for placental and blood vessel function. The most significant SNP in this cluster, rs5960 (p<6×10^−6^) is a synonymous variant in the clotting factor *F10*. Two other members of the coagulation cascade, *F7* and *PROZ*, are also in the top associated region. However, we detected no difference of PROZ levels in maternal or umbilical cord plasma.

**Limitations, reasons for caution:** Our genome-wide association analysis (GWAS) was limited by a small sample size and lack of functional follow up. Our ELISA was limited to post-natal blood sampling (only samples collected immediately after birth). But, despite a small sample size, our family based GWAS design permits identification of novel significant and suggestive associations with preeclampsia. Further longitudinal studies could analyze clotting factor levels and activity in other pregnant cohorts in Peru to assess the impact of thrombosis in preeclampsia risk among Andean highlanders.

**Wider implications of the findings:** These findings support previous evidence suggesting that coagulation plays an important role in the pathology of preeclampsia and potentially underlies susceptibility to other pregnancy disorders exacerbated at high altitudes. This discovery of a novel association related to a functional pathway relevant to pregnancy biology in an understudied population of Native American origin demonstrates the increased power of family-based study design and underscores the importance of conducting genetic research in diverse populations.

**Study funding/competing interest(s):** This work was supported in part by the National Science Foundation (NSF) Graduate Research Fellowship Program Grant No. DGE–1147470 awarded to K.M.B.R. (fellow no. 2014187481); NSF SBE Postdoctoral Research Fellowship Award No. 1711982 awarded to M.N.C.; an A.P. Giannini Foundation postdoctoral fellowship, a Stanford Child Health Research Institute postdoctoral award, and a Stanford Dean’s Postdoctoral Fellowship awarded to E.T.Z.; the Chan Zuckerberg Biohub Investigator Award to C.D.B; a Burroughs Welcome Prematurity Initiative Award to J.C.B.; the George Rosenkranz Prize for Health Care Research in Developing Countries, and the International Center for Genetic Engineering and Biotechnology (ICGEB, Italy) grant CRP/ MEX15-04_EC, and Mexico’s CONACYT grant FONCICYT/50/2016, each awarded to A.M.E. Further funding was provided by the Sandler Family Foundation, the American Asthma Foundation, the RWJF Amos Medical Faculty Development Program, Harry Wm. and Diana V. Hind Distinguished Professor in Pharmaceutical Sciences II, National Institutes of Health, National Heart, Lung, and Blood Institute Awards R01HL117004, R01HL128439, R01HL135156, R01HL141992, National Institute of Health and Environmental Health Sciences Awards R01ES015794, R21ES24844, the National Institute on Minority Health and Health Disparities Awards R01MD010443, and R56MD013312, and the National Human Genome Research Institute Award U01HG009080, each awarded to E.G.B. Author J.W.C. is currently a full-time employee at Genentech, Inc. and hold stocks in Roche Holding AG. Author E.G.B. reports grants from the National Institute of Health, Lung, Blood Institute, the National Institute of Health, General Medical Sciences, the National Institute on Minority Health and Health Disparities, the Tobacco-Related Disease Research Program, the Food and Drug Administration, and the Sandler Family Foundation, during the conduct of the study.

**Trial registration number:** N/A

*for MESH terms see PubMed at http://www.ncbi.nlm.nih.gov/pubmed/

## Introduction

Preeclampsia is a hypertensive disorder of pregnancy that is a leading cause of morbidity and mortality for mothers and infants worldwide. The disorder complicates 5-7% of global pregnancies, causes nearly 40% of all premature births, and is associated with 10-15% of all maternal deaths (Duley, 2009, Rana et al., 2019, Valenzuela et al., 2012). This morbidity is even higher in developing countries and among communities with limited access to healthcare (Osungbade and Ige, 2011). Despite posing a significant global disease burden, the heterogeneity of preeclampsia has posed a major challenge for understanding its etiology and genetic basis (Phipps et al., 2019, Valenzuela et al., 2012).

Clinical and pathological research suggests a major role for the placenta in preeclampsia, where shallow invasion of fetal cells into the maternal endometrium results in insufficient remodeling of the maternal vasculature (Yong et al., 2018). While it roots in early placental development, preeclampsia is usually not detected until the third trimester of pregnancy (>20 weeks gestation), when it is identified by a sudden onset of hypertension and signs of organ damage, typically proteinuria (excess protein in the urine). The severity of preeclampsia is determined by gestational age at onset, as well as the magnitude of hypertension and organ damage (American College of Obstetricians and Gynecologists, 2013). The disorder is known to be heritable with multicomponent risk determined by maternal, fetal, and paternal factors (McGinnis et al., 2017, Pappa et al., 2011, Phipps et al., 2019, Valenzuela et al., 2012). Other risk factors include family history (Boyd et al., 2013, Cincotta and Brennecke, 1998), socioeconomic status (Silva et al., 2008) and chronic hypertension or diabetes (Rana, et al., 2019). Residence at high altitudes above 2,500 meters (m) also contributes considerably to risk of developing preeclampsia (Zamudio, 2007).

Residence at high altitudes increases the risk for preeclampsia and other hypertensive pregnancy disorders at least two to threefold (Moore et al., 2011). For example, Bolivian communities living at 3,500 m altitude have an incidence of preeclampsia of up to 20% (Keyes et al., 2003), about three times higher than the world average (Abalos et al., 2013). In neighboring Peru, preeclampsia complicates up to 22% of all pregnancies and is the second leading cause of maternal deaths (Gil Cipirán, 2017, Guevara Ríos and Meza Santibáñez, 2014). Due to this high incidence, highland pregnancy studies have been proposed as a natural experiment to elucidate genetic factors involved in preeclampsia and other hypertensive pregnancy complications (Moore et al., 1982, Moore et al., 2004, Palmer et al., 1999, Tissot van Patot et al., 2009, Zamudio, 2007). Native Andean populations are of particular interest for this research due to their unique physiological adaptations to chronic high-altitude hypoxia, such as enhanced pulmonary volumes and elevated blood hemoglobin concentrations (Bigham et al., 2013). Candidate genes involved in these adaptations include EGLN1, NOS2 and the hypoxia-inducible factor 1 (HIF1) pathway, among others (Beall, 2014, Bigham, et al., 2013).

Previous research has found that Highland Andean ancestry and long term, multi-generational residence at altitude are associated with lower rates of hypoxia induced pregnancy complications among high altitude resident women (Julian et al., 2009, Moore, et al., 2011, Moore, et al., 2004). Because preeclampsia risk increases with altitude (Palmer, et al., 1999), these findings suggest that Andeans with Native American ancestry may carry rare adaptive variants or a unique repertoire of genetic risk factors for preeclampsia—distinct from other populations previously studied (Michita et al., 2018). Characterizing fine-scale ancestry and genetic structure patterns in native Andeans may uncover preeclampsia relevant genetic variation found at higher frequencies due to selection for altitude adaptation (Bigham and Lee, 2014, Tishkoff, 2015).

To this end, here we analyze genotype data from a large cohort of preeclamptic Andean families from Puno, Peru (Figure 1A). This city, located at 3,830 m altitude, has a population with one of the highest incidences of preeclampsia and associated maternal mortality in the world (Bristol, 2009, Gil Cipirán, 2017). Our work takes a comprehensive approach to the genetic study of preeclampsia in a population adapted to high-altitude by employing a family-study design within a case-control cohort. This enables identification of genetic regions that influence preeclampsia considering each of the family members that affect disease risk—mothers, fathers, and offspring—unlike most genome-wide studies focused on pregnancy disorders which tend to solely include maternal or fetal genomes (Williams and Broughton Pipkin, 2011). We also aim to understand the role of ancestry-related susceptibility in this disorder by characterizing genetic diversity and admixture patterns in the Puno cohort. Additionally, because preeclampsia presents in a spectrum of severity based on gestational age, organ damage, and hypertension, we take advantage of extensive cohort phenotyping to study associations of genetic variants with disease severity. Our findings have implications for general understanding of preeclampsia, and human pregnancy hypertensive disorders more broadly, while also shedding light on the genetic factors that underlie human adaptations for successful reproduction at high altitudes.

**Figure 1.**
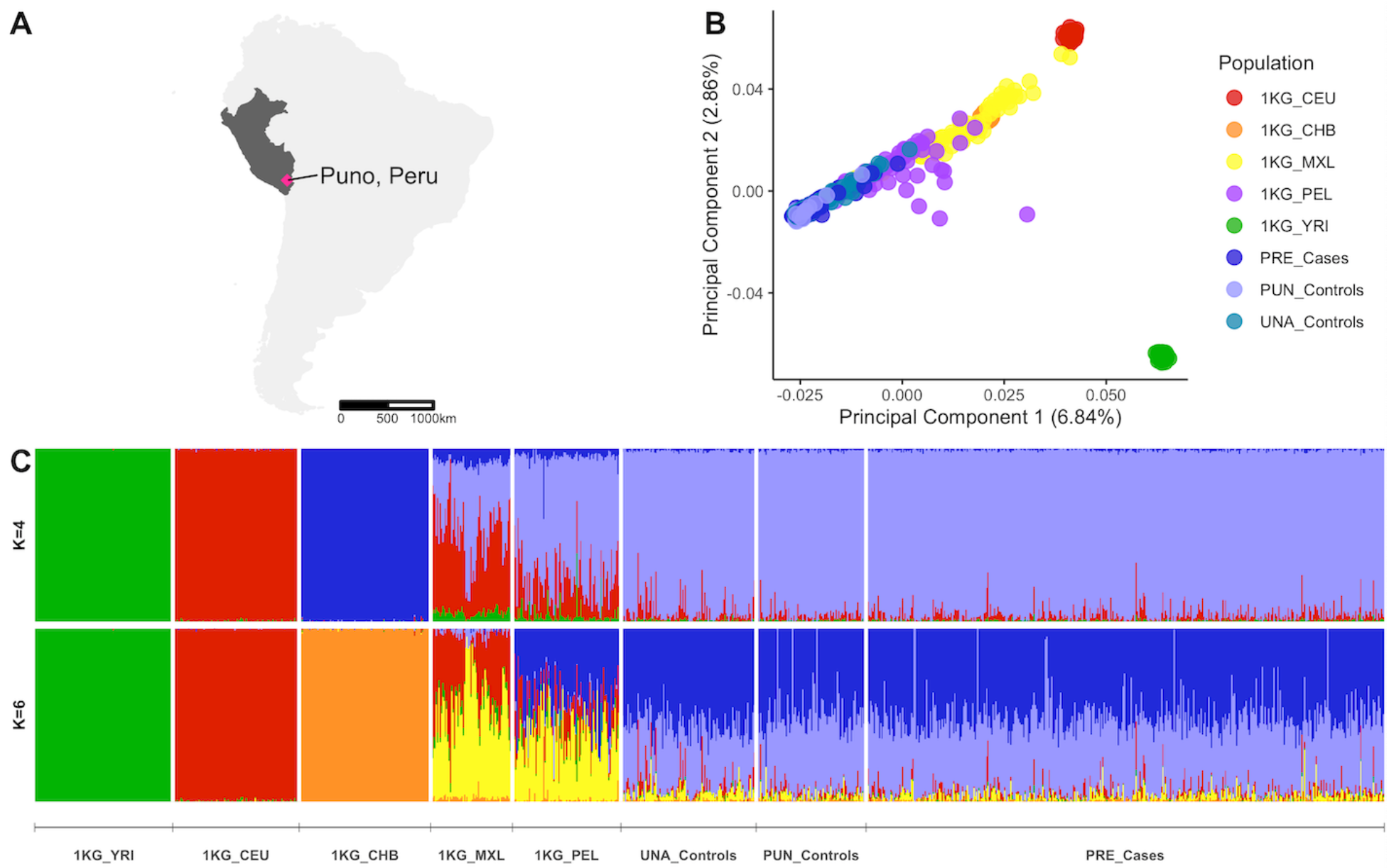
Location and population structure of the Puno preeclampsia cohort. A) Approximate location of Puno, Peru. B) Principal components analysis including PRE cases, PUN and UNA controls, and five continental reference populations from the 1000 Genomes. C) ADMIXTURE analysis results showing unsupervised clustering models assuming K=4 and K=6. At K=6 a Puno-specific sub-continental ancestry component not shared with 1000 Genomes Peruvians from Lima appears in the Puno cohort (shown in light blue).

## Materials & Methods

### Puno cohort

Preeclamptic families (PRE) were recruited between 2011 and 2016 in the Puno regional hospital *(Hospital Regional Manuel Nuñez Butrón*) after their preeclampsia diagnosis. Expecting parents (mothers and fathers) had to be at least 18 years of age and report at least two generations of parents from Puno or nearby Andean regions. Recruited families and subjects included 136 trios (mother, father, and fetal umbilical cord), 197 duos (190 mother and fetal umbilical cord duos, and 7 mother and father pairs), and 14 singletons (mother or umbilical only). 100 healthy same-population control families from Puno (PUN) were also recruited at the hospital at their time of admission for labor. These included 4 trios and 96 duos (mother and fetal umbilical cord). Lastly, 110 unrelated population controls were recruited at the local university, *Universidad Nacional del Altiplano* (UNA) in Puno. In total, 1,129 samples were collected, including 815 PRE cases, 204 PUN and 110 UNA controls (Supplementary Table 1).

### Ethical approval

All participants were recruited with informed consent and with approval by the Stanford University Institutional Review Board eProtocols 20782 (Investigating the Genetic Basis of Preeclampsia in Populations Adapted to High Altitude) and 20839 (Population and Functional Genomics of the Americas). Local IRB approvals were provided by the ethics committee at the Manuel Nuñez Butrón Regional Hospital (01541-11-UADI-HR“MNB”-RED-PUNO) and the Peruvian National Institute of Health (213-2011-CIEI/INS).

### Phenotypic data

Preeclampsia was defined as new onset of hypertension with presence of proteinuria in urine after 20 weeks of gestation. Hypertension was defined as systolic blood pressure 30 mmHg higher than basal level, and diastolic blood pressure at least 15 mmHg higher over basal level. If no prior blood pressure measurements were available, average basal levels were used as prior (85/55 mmHg). Note that measured basal arterial pressure levels in pregnant women in Puno are around 80/50 – 90/60 mmHg (systolic/diastolic), much lower than the U.S. standards, possibly due to altitude adaptation (Segura-Vega, 2019). Proteinuria levels were confirmed to be at least 30mg/dL by dipstick in two tests 24 hours apart. Severity of preeclampsia was defined by the attending physician and categorized into mild or severe. Gestational time was self-reported by the mother (by date of last menstrual period: LMP) or determined by the neonate Capurro test.

### Blood and tissue collection

Whole blood from the mothers was collected within a few hours post-partum by venipuncture into EDTA tubes and frozen at −20C. Umbilical cord blood was collected by venipuncture following clamping of the cord immediately after delivery. Paternal blood, and blood from UNA controls, was obtained upon consent. For plasma, EDTA tubes were spun within 60min of collection at 1,200g for 10min in a tabletop centrifuge. Separated plasma was transferred to Eppendorf tubes, spun again under the same conditions for better purity, then stored at −20C in cryovials.

### Genotypic data

DNA was obtained from whole blood with the Promega (USA) Wizard ® Genomic DNA Purification Kit following manufacturer’s instructions. DNA extracts were initially quantified with the Nanodrop. DNA content and quality were further assessed through quantification with the Qubit® Broad Range Assay and by visualizing on a 1% agarose gel, respectively. Samples that had both >10 ng/uL of DNA concentration and visible bands on the gel were selected for genotyping. Genotype data at over 800,000 sites across the genome were generated with the Affymetrix (USA) Axiom Genome-wide LAT 1 array for 950 samples in two batches. Batch 1 was genotyped in February 2014 at the University of California San Francisco, Gladstone Genomics Core in Mission Bay, San Francisco, CA. This batch included 360 PRE, 10 PUN and 110 UNA individuals (n=480). A total of 813,366 variants were successfully genotyped with Batch 1. Batch 2 was genotyped in November 2018 at Affymetrix Research Services Laboratories, Thermo Fisher Scientific in Santa Clara, CA. This batch included 324 PRE and 146 PUN individuals (n=470), as well as 10 controls added by the genotyping facility. Three samples failed the genotyping facility filtering metrics, therefore a total of 477 samples and 818,154 variants were successfully genotyped with Batch 2.

### Quality control

#### Batch 1 data

The genotyping facility performed a first round of QC restricting the raw dataset to 713,709 recommended SNPs that passed filtering thresholds for heterozygous strength offset, cluster resolution, off-target variants, call rate and genotype quality. We further removed 42 variants with duplicate marker names and flipped 21 SNPs to the forward strand using snpflip (https://github.com/biocore-ntnu/snpflip) and Plink v1.9 (Chang et al., 2015). We revised that all variants had physical positions in the NCBI Build GRCh37 human reference (hg19 assembly). After QC, Batch 1 dataset included 713,667 biallelic SNPs and 480 individuals.

#### Batch 2 data

We removed 214 variants with duplicate marker names, 4,233 structural variants and 540 variants with no physical position in the NCBI Build GRCh37 human reference. 64 SNPs were flipped to the forward strand as above. Additionally, we followed the genotyping facility recommendations to restrict this dataset to 777,946 recommended SNPs that passed filtering thresholds for cluster resolution, off-target variants, call rate and genotype quality. The 10 genotyping controls were also removed. After QC, Batch 2 dataset included 777,946 biallelic SNPs and 467 individuals.

#### Batch 1 and 2 merge

We intersected Batch 1 and 2 datasets at overlapping sites using Plink v1.9. The merged dataset contained 689,528 SNPs and 947 individuals. Using Plink, we removed 1,438 SNPs with genotype missing call frequency >5% (flag: --geno 0.05) and 183,054 SNPs with minor allele frequency (MAF) <0.5% (flag: --maf 0.005). We also excluded two individuals with missing call frequency <10% (flag: --mind 0.1). 561 SNPs failing Hardy-Weinberg equilibrium at 10e-10 were also excluded. We next filtered our dataset for families with excess Mendelian errors, cryptic relatedness, and duplicate samples (see Supplementary Table 2 for list of individuals assigned as unrelated after pedigree revision). 31 individuals were removed, and 56 pedigrees were updated. Chromosomal sex was estimated and sex misassignments were corrected for 176 individuals whose biological sex was either not recorded or incorrectly recorded during data collection. After QC, the merged Batch 1 + 2 dataset included 504,475 genome wide SNPs and 914 individuals (Supplementary Figure 1).

### Batch effect correction

We tested for batch effects by calculating principal components analysis in Plink after filtering the dataset for linkage disequilibrium and removing related offspring (flags: --indep-pairwise 100 10 0.1, --pca). We initially identified a strong batch effect with the top principal components statistically significantly associated with batch *(P<0.05)* (Supplementary Figure 2). To correct this effect, we conducted an additional round of site and sample-specific filtering. We removed symmetrical SNPs (AT, CG), excluded all sites not included in the “Best and Recommended” list provided by Affymetrix for this array, and filtered sites with genotype missingness <5% and MAF >0.5%. Additionally, we removed individuals with excess heterozygosity (outliers >4SD), duplicate individuals and individuals with cryptic or unexpected relatedness. In total, 65,161 SNPs and 31 individuals were removed. We repeated the principal components calculation as above on the filtered dataset and found no statistically significant association between batch and the top principal components (Supplementary Figure 2). The final dataset after batch effect correction included 439,314 genome wide SNPs and 883 individuals.

### Population structure

We intersected our dataset with reference panels including five populations from 1000 Genomes (1KG) Phase 3: Yoruba from Ibadan, Nigeria (YRI), Utah residents with Northern and Western European ancestry (CEU), Han Chinese from Beijing, China (CHB), Mexican Americans from Los Angeles, USA (MXL) and Peruvians from Lima, Peru (PEL). After merging, we removed offspring and related individuals, restricted to autosomes and re-applied quality filters. The filtered, merged dataset consisted of 422,224 variants and 1,057 individuals. The unsupervised clustering algorithm ADMIXTURE (Alexander et al., 2009) was run on this dataset to explore global patterns of population structure. As recommended by the ADMIXTURE manual, the input data was LD pruned using Plink (flag: --indep-pairwise 50 10 0.1). After LD pruning, 45,496 variants remained for analysis. Ten ancestral clusters (K=2 through K=10) were tested and the best fit model was selected after examining cross-validation errors. To account for possible convergence variation, we performed 10 additional runs using different random seeds per run and estimated parameter standard errors using 200 bootstrap replicates per run. ADMIXTURE results were plotted with the R pophelper package (Francis, 2017). Principal components analysis (PCA) was applied to the LD pruned dataset using EIGENSOFT v7.2.1 (Patterson et al., 2006) and plots were generated using the ggplot2 package in R v4.0.3 (R Core Team, 2018, Wickham, 2016).

### Phasing and local ancestry estimation

We used RFMix v1.5.4 (Maples et al., 2013) to determine genome wide local ancestry proportions for the Puno cohort founders, assuming a model of K=3 ancestral populations. The choice of K=3 reference populations was informed by the ADMIXTURE results. The reference panel included 108 YRI and 94 CEU individuals from 1000 Genomes Phase 3, and 94 native individuals from Mexico (30 Mixe, 15 Zapotec, 49 Nahua) genotyped as part of the GALA II study (Galanter et al., 2014). These reference samples were used as proxies for African, European, and Native American ancestral source populations, respectively. After merging, the analysis ready dataset consisted of 420,105 overlapping variants and 899 individuals. The data were phased with SHAPEIT2 (O’Connell et al., 2014). RFMix was run with default parameters and EM=2 iterations. Ancestry call cutoffs were determined with a 0.9 posterior probability threshold as recommended in (Kidd et al., 2012).

### Ancestry proportions analysis

We tested for significant differences in proportions of Native American, European, and African ancestry components between PRE cases, PUN and UNA controls. We applied the Wilcoxon signed ranks test in R v3.5.1 (pairwise.wilcox.test function) with Bonferoni correction for multiple testing. This non-parametric test assesses whether significant differences exist between two distributions (Moore et al., 2009). Our null hypothesis was that the distribution of each ancestry proportion was identical between PRE cases, PUN and UNA controls.

### Statistical analysis of clinical phenotypes

We assessed batch bias of clinical phenotypes and correlation with each other by statistical analysis in R v3.4.0 (R Core Team, 2018). The following dichotomous phenotypes were tested for batch association with a chi squared test: severity of diagnosis (mild or severe), proteinuria (+/++ or +++), parity (nulliparous or more than one birth), sex of newborn and mode of delivery (vaginal or C-section). The following continuous phenotypes were tested for batch association by t-test: gestational time measured by the mother (date of last menstrual period, or LMP) and by the fetus (Capurro test), neonate weight, systolic and diastolic blood pressure measurements, and maternal age.

### Transmission-disequilibrium test (TDT) and parent of origin (POO)

Leveraging the trio family structure, we applied the transmission disequilibrium test (TDT) and parent-of-origin (TDT-POO) test on all 87 parent-offspring case trios (preeclamptic families with offspring) in Plink v1.9 using the --tdt flag, with and without the ‘poo’ modifier. Variants were then filtered by MAF > 0.05 within the analyzed cohort. The TDT test assumes Mendelian rules for transmission of alleles and tests if the queried allele is being transmitted/untransmitted disproportionately from parents to the affected offspring population (Purcell et al., 2007, Purcell et al., 2005). The POO analysis is part of TDT, and separately queries transmission from each parent individually to assess paternal or maternal specific transmission. This test self-corrects for covariate effects by treating each trio as a separate unit.

### GWAS for case-control association

Puno cohort individuals were divided into offspring and mothers for two separate case-control GWAS analyses using logistic regression in Plink (flag: --logistic) with the first 3 PCs and sequencing batch as covariates. The analysis on the mothers includes 254 PRE and 70 PUN controls. The offspring analysis includes 225 PRE cases and 60 PUN controls. These analyses included individuals in trios, duos, and singletons. Variants were filtered by MAF > 0.05 within the analyzed cohort.

### GWAS in additional phenotypes

Multiple phenotypes measured and captured in the recruited patient’s medical history allow for testing of additional genetic associations. We performed additional genome-wide association analyses of endophenotypes in the PRE mothers (N=254) and offspring (N=225), separately. These analyses included individuals in trios, duos, and singletons. The endophenotypes tested for each were: (1) gestational age, maternal measurement; (2) gestational age, fetal measurement; (3) diastolic blood pressure at diagnosis of preeclampsia; (4) systolic blood pressure at diagnosis of preeclampsia; (5) proteinuria at diagnosis and (6) severity of diagnosis. The first four were treated as continuous variables and analyzed by linear regression in Plink (flag: --linear). Proteinuria and severity of diagnosis were dichotomous variables analyzed in Plink by logistic regression (flag: --logistic), with proteinuria reduced to + and ++ vs. +++. Genotyping batch was included as a discrete covariate and the first 3 PCs as continuous covariates. Several of these analyses included less individuals due to missing data. Specifically, GWAS with systolic and diastolic blood pressure included 253 PRE mothers and 224 PRE offspring, and GWAS with maternal measurement of gestational age included 252 PRE mothers and 223 PRE offspring.

### GWAS data visualization

All genome-wide analyses were filtered by MAF >= 0.05 within the analyzed cohorts and visualized by Manhattan plots using the qqman R package v0.1.4 (Turner, 2017). QQ plots were generated with the same package to confirm no effects from population structure or other confounders. Regions of interest were selected if they met two criteria: (1) p-value (p<10E-4 in most cases—unless specified in the results section) and (2) the presence of nearby associated SNPs forming a skyscraper-like structure in the Manhattan plot. Top SNPs in these regions were selected, and their genomic regions plotted using LocusZoom (Pruim et al., 2010). Maps displaying the geographic distribution of candidate associated variants were produced using the Geography of Genetic Variants (GGV) browser (Marcus and Novembre, 2017).

### Capture sequencing

We conducted fine mapping of potential causal variants in a subset of families genotyped in Batch 1 previous to Batch 2 genotyping. Preliminary data obtained from Batch 1 genotypes were analyzed using standard family-based TDT on Plink for preeclampsia associations (as above), and regression analysis on secondary phenotypes was conducted using linear mixed models in GTCA (Yang et al., 2011) (flag: –mlma-loco). Based on these preliminary results, we designed a target capture assay including windows around top hits for preeclampsia and secondary phenotypes, as well as several genes previously suggested to be associated with preeclampsia in the GWAS catalog (release 2.0.5) (Buniello et al., 2019). The total capture size was approximately 10Mb (Supplementary File 1).

We next selected families from Batch 1 with the strongest associations on the preliminary TDT analysis (n=86 individuals, Supplementary Table 1). Genomic DNA from 86 individuals (Supplementary Figure 3) was fragmented by mechanical shearing (Covaris) and prepared using the KAPA Hyperprep library preparation kit (Kapa Biosystems, now part of Roche, Switzerland). DNA capture was performed on the libraries using the Agilent (USA) SureSelect platform following manufacturer’s instructions. Paired-end sequencing of captured libraries was performed on the Illumina NextSeq. Sequence data were analyzed through a standard FASTQC-BWA-GATK pipeline following guidelines as described in (Koboldt, 2020). We then performed the same GWAS analyses listed above (TDT test for the preeclampsia phenotype and linear regressions for continuous phenotypes) in the captured regions in a limited set of individuals: 25 trios, 4 duos (3 mother-offspring, 1 father-offspring) and 3 singletons (1 offspring and 2 mothers). Candidate loci identified in these analyses were individually merged and annotated with ANNOVAR (Yang and Wang, 2015) and overlapped with GTEx single-tissue cis-eQTL data (version V6p) from the online database (https://gtexportal.org/home/datasets) to find relevant GTEx annotations in our data set (Carithers et al., 2015, Carithers and Moore, 2015).

### ProZ ELISA

ProZ levels in post-partum maternal and cord blood plasma were assayed using the human-ProZ ELISA kit from MyBioSource (USA, Cat. No. MBS765710), following manufacturer instructions. Maternal and fetal plasma samples were diluted at 1:400 in sample diluent and all washes were performed manually with a multichannel pipet. Final optical density absorbance at 450nm was read using the Bio Rad (USA) iMark^TM^ Microplate Absorbance reader. A 4-Parameter curve fit was applied to the standards, and the resulting equation was used to calculate concentration in the experimental samples. Boxplots and t-tests were done in R v3.4.0 (R Core Team, 2018).

## Results

We obtained blood samples and maternal clinical records from consented families at the *Hospital Regional Manuel Nuñez Butrón*, and blood alone from individuals recruited at the *Universidad Nacional del Altiplano*. At the time of recruitment, mothers from case families (labeled PRE throughout this study) were at hospital experiencing pregnancy with a preeclampsia diagnosis, defined as hypertension and proteinuria after 20 weeks of gestation. It is important to note that basal blood pressure in this population is lower than in the U.S., and hypertensive levels can be as low as 110/65 mmHg, compared to 140/90mmHg in U.S. guidelines. Rather than based on a cutoff, hypertension was defined as a systolic measurement 30 mmHg higher than basal and diastolic at least 15 mmHg higher than basal for each individual (see Materials & Methods for more details). For consistency, and to control for other hypertensive complications of pregnancy, we included proteinuria in the diagnosis, despite this factor not being currently required in many diagnostic guidelines (American College of Obstetricians and Gynecologists, 2020).

Mothers from control families (labelled PUN) were experiencing a pregnancy without complications at time of hospital recruitment. 88 PRE families and two PUN families were collected as complete trios—including both biological parents and offspring; the rest are duos (one parent and offspring) and single individuals (mothers) (Table I). Overall, the Puno cohort collected for this study includes 815 individuals from the PRE group, 204 from the hospital control group (PUN), and 110 from the university (UNA) as ‘population controls. We extracted DNA from blood and genotyped PRE, PUN and UNA individuals in two batches on the Affymetrix Axiom LAT array. Our final dataset after quality filtering included 439,314 genome wide SNPs and 883 individuals (see Table I and Supplementary Table 1 for breakdown of PRE, PUN and UNA).

**Table I.** All individuals genotyped by group (case/control) and batch after QC filtering.

| Family Category<sup>1</sup> | PUN and UNA Controls |  |  |  | PRE Cases |  |  |  | Total No. of Individuals |
| --- | --- | --- | --- | --- | --- | --- | --- | --- | --- |
|  | Batch 1 individuals | Batch 2 individuals | Batch 1+2 individuals | Batch 1+2 family units | Batch 1 individuals | Batch 2 individuals | Batch 1+2 individuals | Batch 1+2 family units |  |
| **Trios <sup>2</sup>**<br>**(M+F+UC)** | **0** | **6** | **6** | **2** | **241** | **21** | **262** | **88<sup>2</sup>** | **268** |
| **Duos** | **10** | **106** | **116** | **58** | **110** | **192** | **302** | **151** | **418** |
| *M+UC* | *8* | *106* | *114* | *57* | *66* | *188* | *254* | *127* | *368* |
| *P+UC* | *2* | *0* | *2* | *1* | *18* | *2* | *20* | *10* | *22* |
| *M+F* | *0* | *0* | *0* | *0* | *26* | *2* | *28* | *14* | *28* |
| **Singletons <sup>3</sup>** | **106** | **24** | **130** | **--** | **19** | **48** | **67** | **--** | **197** |
| **All individuals** | **116** | **136** | **252** | **60** | **370** | **261** | **631** | **239** | **883** |

<sup>1</sup>M= Mother, F=Father, UC=Umbilical cord.
<sup>2</sup> Includes one trio with two offspring from family PRE061.
<sup>3</sup> Includes individuals coded as UNR (unrelated and no longer connected to medical records), UNA (university controls), and some individuals collected as part of PRE/PUN (still connected to medical records).

### Puno individuals have high proportions of Native American ancestry

We sought to understand the demographic history of our test population by characterizing patterns of genetic diversity and population structure in the Puno study cohort. To this end we intersected the entirety of the Puno cohort dataset (883 individuals) with a reference panel including five continental populations from the 1000 Genomes (1KG) Project Phase 3 panel: Yorubans (YRI), Europeans (CEU), Mexicans (MXL), Han Chinese (CHB) and Peruvians from Lima (PEL). Using principal component (PC) analysis, we find that individuals from Puno (either PRE, PUN, UNA) cluster together in PC space, and are distributed in a clinal pattern alongside Peruvians from Lima who have high proportions of Native American ancestry (Figure 1B, Supplementary Figure 4).

We next investigated admixture patterns in the Puno population with the goal of characterizing proportions of Native versus non-Native genomic ancestry. Using the clustering algorithm ADMIXTURE (Alexander, et al., 2009), we explored unsupervised models assuming K=2 through K=10 ancestral clusters (Supplementary Figure 5). Cross-validation errors for each K cluster are shown in Supplementary Figure 6. At K=4, we observe a clear separation of continental-scale ancestry components. We find that Puno individuals have large proportions of Native American ancestry and small proportions of European ancestry, represented by blue and red in Figure 1C, respectively. At the best fit model of K=6, ADMIXTURE analysis finds substructure within the Native American ancestry component of the Puno cohort. Specifically, we observe a Puno-specific ancestry component (shown in light blue in Figure 1C) which is not present within the Native American ancestry components of 1KG Mexican and Peruvian individuals. This substructure may derive from an Andean specific ancestry component that has been previously identified among Indigenous and mestizo communities from the Andean Highlands (Barbieri et al., 2019, Harris et al., 2018). Overall, we find that individuals in the Puno cohort are predominantly of Native American ancestry (95.7% on average) and have low levels of non-Native American admixture (approximately 4.2% on average; Supplementary Table 3). We further find that the Puno population carries a Highland-specific Native American sub-continental ancestry component, as noted in previous work (Barbieri, et al., 2019, Harris, et al., 2018).

Finally, we tested for significant differences in ancestry proportions between cases (PRE) and controls (PUN, UNA) in the Puno cohort. Guided by the findings of the ADMIXTURE analysis, we used RFMix to determine local ancestry proportions in the Puno cohort assuming a model of K=3 ancestral components. We next extrapolated average ancestry proportions per individual from the RFMix local ancestry calls (Supplementary Tables 4-5). The results of this estimation further confirm the predominantly Native American ancestry background and highlight the small proportion of European admixture present in our sample. We next performed a Wilcoxon rank test to contrast ancestry proportions between PRE, PUN and UNA. This test identified a small but significant difference in European ancestry proportions between PRE and UNA but found no significant differences in Native American or African ancestry proportions (Supplementary Figure 7, Supplementary Table 6). Overall, UNA individuals have slightly higher proportions of European ancestry than PRE and PUN individuals. However, proportions of Native American ancestry are not significantly different between cases (PRE) and controls (PUN, UNA). These findings support the results of the ADMIXTURE analysis and further underscore the primarily Native American ancestry background of the Puno cohort.

### Family-based analysis reveals association of a cluster of clotting factor genes (*PROZ*, *F7*, *F10*) with preeclampsia

Next, we sought to identify genetic loci associated with the risk of preeclampsia in this highly susceptible population adapted to the hypoxic conditions of the Andean Highlands. As decades of genetic research have shown a role for maternal, paternal and offspring genomes on preeclampsia risk (Galaviz-Hernandez et al., 2018, Gray et al., 2018, Phipps et al., 2019), we collected family trios from 88 cases, as well as duos when trio sampling was not possible (either for lack of consent or due to samples failing genotyping QC), enabling all three genomes to be evaluated. Since preeclampsia is a complex disease with wide ranging phenotypes, we provide summaries of relevant phenotypic data for all case pregnancies organized by batch and in trio cases only (Table II). By statistical comparison, we find that there is moderate batch bias in approximately half of the measured phenotypes (e.g., Batch 2 had significantly more vaginal deliveries than C-sections, when compared to Batch 1, p<0.04), but none likely to influence the analysis when supported by batch correction. In addition to the data shown in Table II, most mothers (>98%) had no history of chronic hypertension or diabetes and all were non-smokers.

**Table II.** Phenotypic characteristics of analyzed case families with preeclampsia (duos and trios). The sum of batch 1 and 2 correspond to the total in the first column. “Trios only” identifies the subset from the total that are in whole trio units (the rest are mother-offspring duos). The last column represents chi-squared or t-test p-values for each phenotype between batches. Significant tests with p<0.05 are identified with an asterisk (*).

|  | **Total** | **Batch 1** | **Batch 2** | **Trios only** | **Batch pval** |
| --- | --- | --- | --- | --- | --- |
| **Diagnosis, *n*** | Mild (n=119) | 63 | 56 | 47 | 0.68 |
|  | Severe (n=106) | 60 | 46 | 41 |  |
| **Mode of delivery, *n*** | C-section (n=92) | 58 | 34 | 42 | 0.04* |
|  | Vaginal (n=132) | 64 | 68 | 45 |  |
| **Maternal age, years**<br>***mean(median)*** | 26.64(26.00) | 26.75(25.00) | 26.51(26.00) | 27.44(26.00) | 0.76 |
| **Gestational age - maternal, weeks**<br>***mean(median)*** | 38.11(39.00) | 37.80(38.00) | 38.48(39.00) | 37.84(38.00) | 0.04* |
| **Gestational age - fetal, weeks**<br>***mean(median)*** | 38.29(39.00) | 38.20(39.00) | 38.39(39.00) | 38.17(39.00) | 0.52 |
| **Newborn weight, grams**<br>***mean(median)*** | 601.3(600.0) | 611.9(605.0) | 569.6(565.0) | 620.6(620.0) | 0.07 |
| **Systolic BP at admission, mmHg**<br>***mean(median)*** | 133.0(130.0) | 131.1(130.0) | 135.3(130.0) | 130.9(130.0) | 0.02* |
| **Diastolic BP at admission, mmHg**<br>***mean(median)*** | 89.82(90.00) | 87.68(90.00) | 92.43(90.00) | 87.1(85.0) | 2.12E-05* |
| **Parity, *n*** | nulliparous (n=128) | 72 | 56 | 45 | 0.68 |
|  | 1 or more (n=97) | 51 | 46 | 43 |  |
| **Sex of newborn, *n*** | female (n=100) | 46 | 54 | 35 | 0.03* |
|  | male (n=125) | 77 | 48 | 53 |  |
| **Proteinuria, *n*** | + (n=124) | 64 | 60 | 49 | 0.38 |
|  | ++/+++ (n=101) | 59 | 42 | 39 |  |

To find genetic linkage between genomic loci and preeclampsia, we first performed a parent-offspring trio GWAS analysis, or transmission-disequilibrium test (TDT), in the 88 affected (PRE) trios. The TDT offers a robust association test of genotype to phenotype in affected families by measuring over-transmission of alleles from heterozygous parents to the offspring. With this analysis, we identified a group of SNPs in linkage disequilibrium (LD) over a cluster of blood clotting factor genes with a high odds ratio for preeclampsia (Figure 2; Table III; Supplementary Figure 8). The most significant SNP in this cluster, rs5960 (OR 3.05, 95% CI 1.841-5.054, p<6×10^−6^; 1000G MAF 0.623), is a synonymous variant in the clotting factor *F10*. Two other members of the coagulation cascade, *F7* and *PROZ*, are also in this region. Another top hit in the TDT, SNP rs553316 (OR 0.339, 95% CI 0.2041-0.5629, p=1.15E-05; 1000G MAF 0.408), is in high LD with rs5960 in 1KG Peruvian populations (R^2^=0.7476) (Machiela and Chanock, 2015). Additionally, rs553316 is annotated in GTEx as an eQTL for *PROZ* on mammary tissue (note that, as of our analysis, no placental or pregnancy blood data were available on GTEx). The global distribution of allele frequencies for rs5960 and rs553316 in 1KG reference populations are shown in Supplementary Figure 9 and noted in Supplementary Table 7.

**Figure 2.**
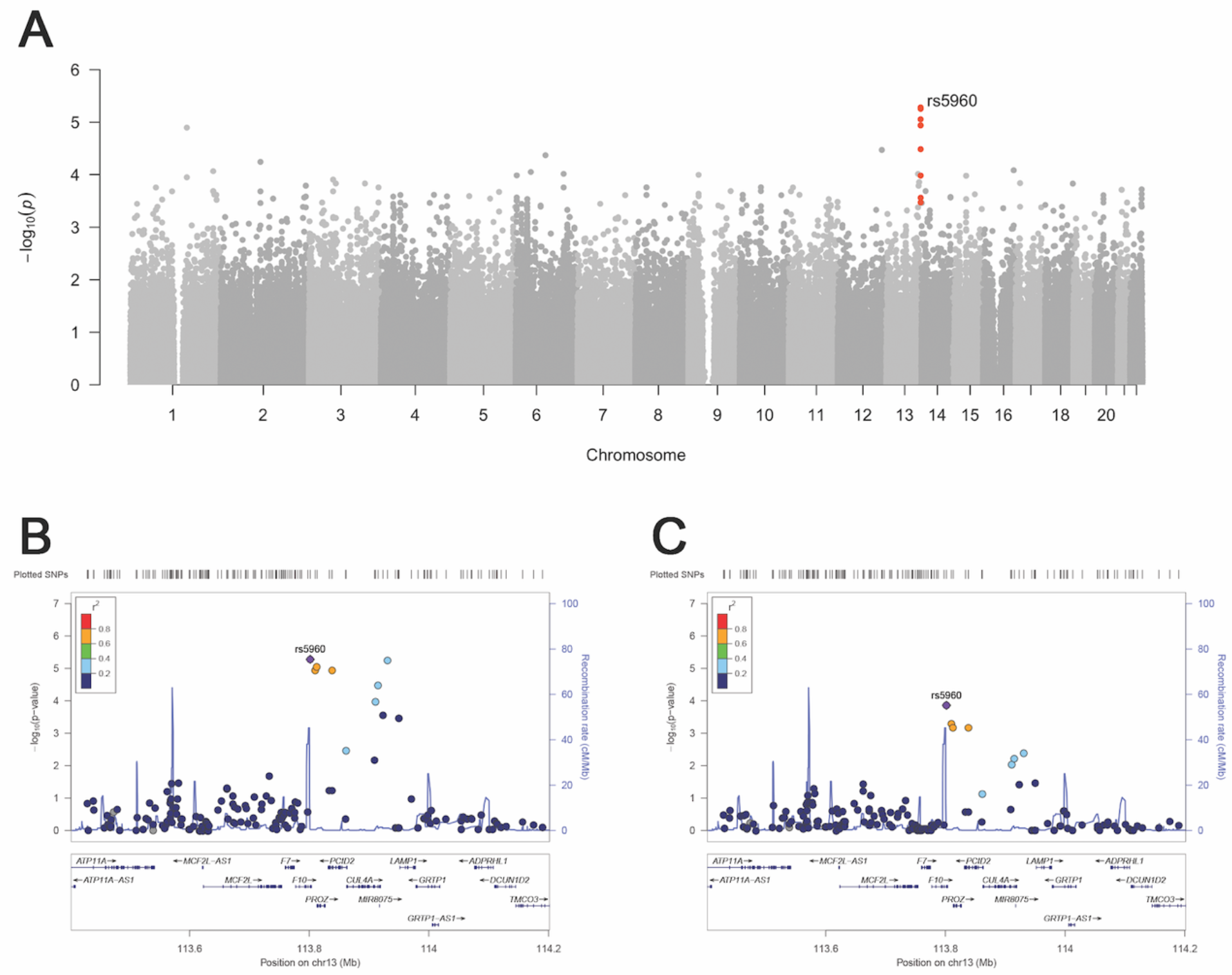
Top associations from trio analyses by TDT and TDT-POO. A) Manhattan plot showing top association with preeclampsia in the offspring genome: SNP rs5960 on chromosome 13 at p<10e-5 suggestive of significance (shown in red). B) Locus Zoom plot depicting the top associated SNP cluster from the TDT on chromosome 13. C) Locus Zoom plot depicting the top paternal region from TDT-POO analysis on chromosome 13.

**Table III.** GWAS statistics and genomic annotations of top hits (P<5×10^−4^) from the TDT.

| Chr | BP | cytoBand | rsID | Ref | Alt | TDT GWAS stats |  |  | Function | Genes in region | Puno MAF |
| --- | --- | --- | --- | --- | --- | --- | --- | --- | --- | --- | --- |
|  |  |  |  |  |  | OR | 95% CI | P |  |  |  |
| 13 | 113801737 | 13q34 | rs5960 | C | T | 3.05 | 1.841-5.054 | 5.23E-06 | exonic | F10 | 0.4929 |
| 13 | 113930853 | 13q34 | rs9549724 | C | T | 0.2963 | 0.1696-0.5176 | 5.58E-06 | intergenic | CUL4A, LAMP1 | 0.4278 |
| 13 | 113812962 | 13q34 | rs2273971 | A | G | 0.3276 | 0.1951-0.55 | 8.81E-06 | upstream | PROZ | 0.499 |
| 13 | 113838015 | 13q34 | rs553316 | G | A | 0.339 | 0.2041-0.5629 | 1.15E-05 | intronic | PCID2 | 0.4827 |
| 13 | 113810186 | 13q34 | rs7335409 | T | C | 2.95 | 1.777-4.899 | 1.15E-05 | intergenic | F10, PROZ | 0.4939 |
| 13 | 113915303 | 13q34 | rs3814260 | G | A | 0.3396 | 0.199-0.5797 | 3.27E-05 | intronic | CUL4A | 0.4236 |
| 2 | 109581319 | 2q12.3 | rs260692 | C | T | 19 | 2.544-141.9 | 5.70E-05 | intronic | EDAR | 0.0551 |
| 1 | 228715705 | 1q42.13 | rs11586639 | G | A | 0.4 | 0.2492-0.6422 | 8.57E-05 | intergenic | BTNL10,<br>MIR7641-2 | 0.4587 |
| 6 | 42252385 | 6p21.1 | rs9471831 | A | G | 0.4242 | 0.2727-0.6601 | 8.88E-05 | intronic | TRERF1 | 0.4562 |
| 13 | 113910926 | 13q34 | rs3861723 | A | G | 0.36 | 0.2101-0.617 | 1.04E-04 | intronic | CUL4A | 0.4228 |
| 1 | 228805855 | 1q42.13 | rs765070 | C | T | 0.4107 | 0.2528-0.6673 | 2.05E-04 | intergenic | DUSP5P1, RHOU | 0.4347 |
| 2 | 109557099 | 2q12.3 | rs260711 | T | C | 9.5 | 2.213-40.78 | 2.08E-04 | intronic | EDAR | 0.05612 |
| 1 | 229076157 | 1q42.13 | rs10916389 | G | A | 0.325 | 0.1738-0.6076 | 2.08E-04 | intergenic | RHOU, MIR4454 | 0.2368 |
| 13 | 113923202 | 13q34 | rs77626225 | A | G | 0.3878 | 0.2283-0.6586 | 2.75E-04 | intergenic | CUL4A, LAMP1 | 0.2307 |
| 13 | 113949751 | 13q34 | rs9549380 | G | A | 0.4 | 0.2382-0.6718 | 3.36E-04 | intergenic | CUL4A, LAMP1 | 0.3201 |
| 2 | 109513601 | 2q12.3 | rs3827760 | A | G | 9 | 2.088-38.79 | 3.47E-04 | exonic | EDAR | 0.05793 |

Given the importance of clotting genes in pregnancy, we sought to complement the genotype analysis by performing deep sequencing of targeted genomic regions surrounding rs5960 in a subset of cohort participants (Supplementary Table 8, Supplementary Figure 3). To fine-map potential causal variants, we repeated the same TDT analysis described above in the fine-mapped individuals and cross-referenced with the GTEx database for expression phenotypes in relevant tissues. This analysis found a strong association of preeclampsia with several eQTLs for *PROZ* (Supplementary Table 9). Other top hits from the genotype TDT that were recapitulated in this analysis include variants in the SLC46A3 and CUL4A genes, also located on chromosome 13 (Supplementary Table 9). Both genes have been previously associated with preeclampsia risk in clinical studies (McGinnis, et al., 2017, Tan et al. 2017). These data suggest that clotting factors on chromosome 13 may play an important role in preeclamptic pregnancies.

Finally, we asked whether this *PROZ* eQTL resulted in differential PROZ protein expression between PRE cases and PUN controls. Since the TDT identifies associated variants in the offspring, we analyzed the umbilical cord plasma of 8 PUN controls and 16 PRE cases by ELISA. In this limited sample, we detected no difference of PROZ levels in umbilical cord plasma (difference in means = 41.550 ug/mL, 95% CI -342.758 to 425.858, p = 0.85) collected after delivery (Supplementary Table 10, Supplementary Figure 10). However, future testing could evaluate PROZ levels in the placenta, where interaction with the maternal environment is more significant to the preeclampsia phenotype than in umbilical cord blood.

### Clotting factor locus shows paternal inheritance

We next examined whether there were loci associated with preeclampsia that were disproportionately inherited either maternally or paternally. To this end, we performed parent-of-origin TDT GWAS in the same 88 trios tested above. This test investigates whether any of the associated SNPs are disproportionately inherited from fathers versus mothers, and vice versa. The most significant SNP from the TDT analysis, rs5960 in *F10*, is suggested to be paternally inherited more often than expected by chance (p=10^−4^, Figure 2, Table IV, Supplementary Figure 11). Other loci show evidence of paternal inheritance, such as rs79278805 (p = 1.77E-04), located within SPAG6 on chromosome 10, and rs9399401 (p=2.76E-04) in ADGRG6/GPR126 on chromosome 6. Similarly, we find several SNPs that show maternal origin bias. The most significant is rs130121 (p=1.91E-04) on chromosome 22 in the FAM19A5/TAFA5 gene, followed by rs10282765 (p=2.39E-04) on chromosome 8 within a ncRNA (Table IV, Supplementary Figures 12-13). Several genes in the vicinity of these SNPs have been implicated in reproduction. SPAG6 is recognized by anti-sperm antibodies and might be involved in infertility (Cooley et al., 2016, Neilson et al., 1999). ADGRG6/GPR126 is a G-coupled protein receptor involved in angiogenesis. It is upregulated in umbilical vein endothelial cells and was found previously to be upregulated in preeclamptic placentas (Cui et al., 2014, Sitras et al., 2009). Overall, these parent-of-origin effects support the hypothesis that maternal and/or paternal bias might contribute to preeclampsia disease.

**Table IV.**
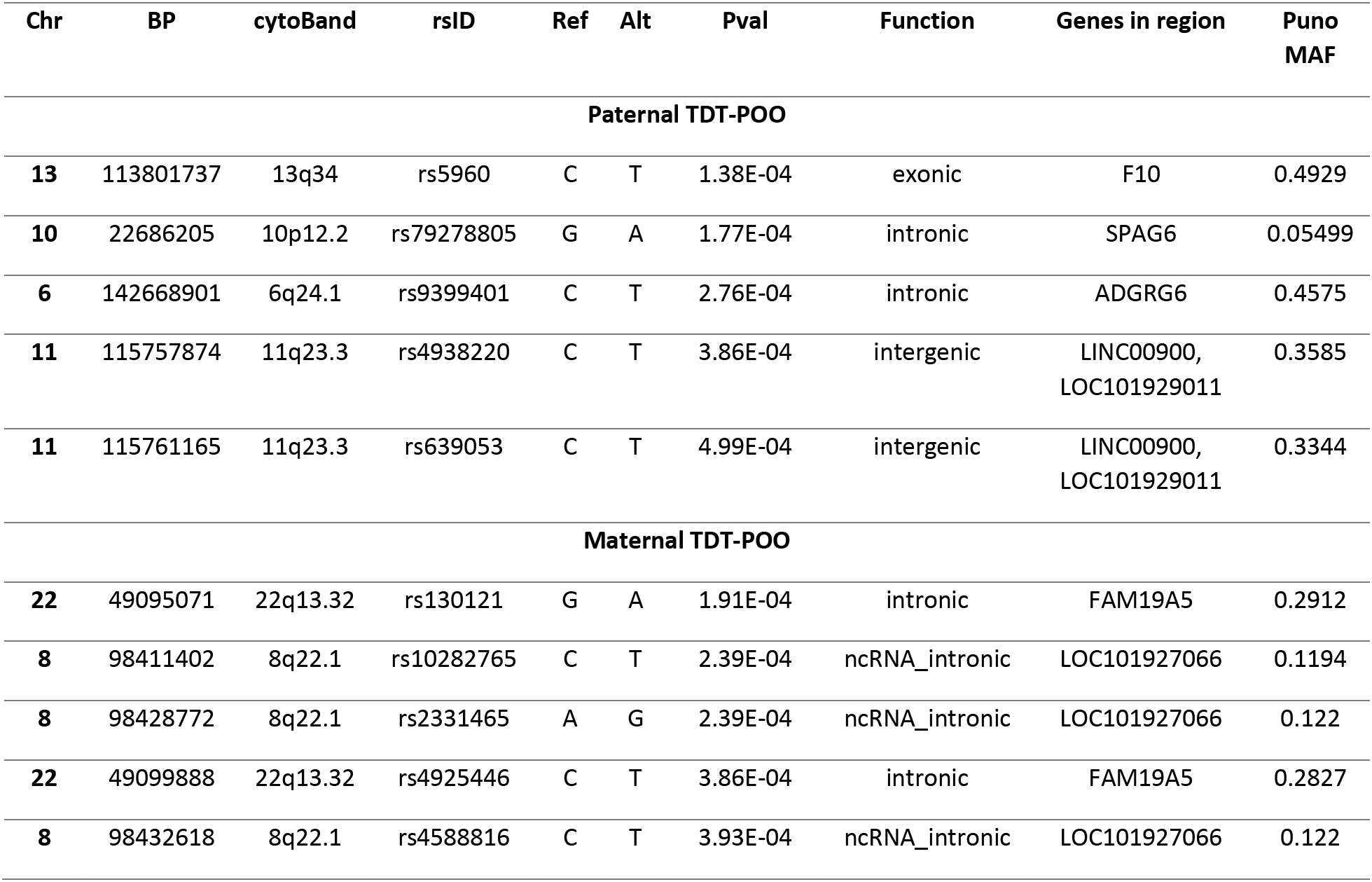
GWAS P values and genomic annotations of top hits (P<5×10^−4^) from the TDT-POO.

**Table V.** Statistics and annotations of the top SNPs (p<5×10^−4^) with biological relevance for preeclampsia of secondary phenotype and case-control GWAS analyses. All SNPs in this table are described in the text (for a complete list of regions at p<5×10^−4^, see supplemental tables. Beta values are reported for linear regressions and odds ratio (OR) for logistic regressions. GA, gestational age; BP, blood pressure.

| GWAS | Chr | BP | cytoBand | rsID | Ref | Alt | GWAS stats |  |  | Function | Genes in region | Puno MAF |
| --- | --- | --- | --- | --- | --- | --- | --- | --- | --- | --- | --- | --- |
|  |  |  |  |  |  |  | BETA/OR | 95% CI | P |  |  |  |
| GA, fetal measurement, maternal genome | 1 | 119404210 | 1p12 | rs952593 | T | C | -1.656 | -2.273 – -1.039 | 3.12E-07 | intergenic | SPAG17, TBX15 | 0.13 |
| GA, maternal measurement, maternal genome | 1 | 119425100 | 1p12 | --- | A | C | -1.439 | -2.013 – -0.8641 | 1.69E-06 | downstream | TBX15 | 0.14 |
|  | 11 | 57203942 | 11q12.1 | rs2581927 | C | T | -2.034 | -2.887 – -1.182 | 4.85E-06 | intergenic | SLC43A3, RTN4RL2 | 0.06 |
| Diastolic BP, maternal genome | 4 | 85200613 | 4q21.23 | rs1874237 | G | A | -4.257 | -5.711 – -2.804 | 2.79E-08 | ncRNA intronic | LOC101928978 | 0.45 |
| Severity, maternal genome | 11 | 94360812 | 11q21 | rs1940640 | T | G | 2.407 | 1.622 – 3.572 | 1.30E-05 | ncRNA intronic | LOC105369438 | 0.43 |
| Proteinuria, maternal genome | 17 | 2028106 | 17p13.3 | rs2760751 | A | G | 2.829 | 1.806 – 4.432 | 5.65E-06 | intronic | SMG6 | 0.29 |
|  | 11 | 94356914 | 11q21 | rs12276362 | C | T | 0.4146 | 0.2795 – 0.6148 | 1.19E-05 | ncRNA intronic | LOC105369438 | 0.49 |
| Systolic BP, fetal genome | 2 | 18099832 | 2p24.2 | rs4553827 | C | T | 7.443 | 4.622 – 10.26 | 5.26E-07 | intronic | KCNS3 | 0.25 |
| Proteinuria, fetal genome | 4 | 5341148 | 4p16.2 | rs62297274 | C | T | 0.3512 | 0.2257 – 0.5465 | 3.51E-06 | intronic | STK32B | 0.49 |
|  | 3 | 25052754 | 3p24.2 | rs4241542 | C | T | 0.2598 | 0.1443 – 0.4678 | 7.04E-06 | intronic | RARB | 0.21 |
| Case-Control, Offspring | 4 | 6671568 | 4p16.1 | rs34360485 | A | G | 3.615 | 2.003 – 6.524 | 1.99E-05 | downstream | LINC02482, S100P | 0.36 |

### Case-control analysis, placental gene S100P is associated with preeclampsia in the offspring

While the TDT identifies preeclampsia risk variants from inheritance analysis, a more common way to test for disease risk variants is to compare cases and controls. The collection of control (PUN) mother-offspring duos allowed us to compare preeclamptic to healthy pregnancies in both the mothers and the offspring. To this end, we performed two case-control GWAS of preeclampsia using Plink (see Materials & Methods): (1) 268 PRE vs. 70 PUN mothers; and (2) 230 PRE and 60 PUN offspring. Several genetic regions showed suggestive association with preeclampsia in both test groups (Supplementary table 11; Supplementary Figures 14-15). The most interesting association was the top SNP in the offspring, rs34360485 on chromosome 4 (p <2E-5, OR 3.615, 95% CI 2.003-6.524, MAF 0.36, (Table V), which contains the placental gene *S100P*. S100P is a calcium-binding protein strongly expressed in the placenta (Zhu et al., 2015) that promotes trophoblast proliferation in culture (Zhou et al., 2016). The global distribution of allele frequencies for rs34360485 in 1KG reference populations is shown in Supplementary Figure 16 and noted in Supplementary table 9.

### Associations of secondary phenotypes reveal loci with roles in placental biology

Preeclampsia is a heterogeneous disease with varying potential markers of severity. For instance, the earlier in gestation preeclampsia occurs, the more severe it is considered to be (Gong et al., 2012, Wojtowicz et al., 2019). Likewise, all the characteristic clinical features associated with preeclampsia (such as proteinuria and elevated blood pressure) can present at varying levels of severity. Harnessing the availability of clinical records for all individuals in the PRE cohort, we next performed GWAS tests on six secondary phenotypes of preeclampsia measured at the time of diagnosis: (1) gestational age, maternal measurement; (2) gestational age, fetal measurement; (3) diastolic blood pressure; (4) systolic blood pressure; (5) proteinuria and (6) severity of diagnosis as stated by the clinician. It is worth clarifying that gestational age (the time of the fetus in the womb) was measured in two different ways throughout the study. The fetal measurement was done by the “Capurro” test, which combines five different measurements in the neonate, while the maternal measurement relies on the date of the mother’s last menstrual period before pregnancy.

To investigate possible genetic associations with secondary phenotypes of preeclampsia, we performed GWAS analyses by logistic and linear regression for each of the six phenotypes in 254 mothers and 225 offspring, separately. In total, we ran 12 GWAS tests. Logistic regression was applied to binary phenotypes (proteinuria and severity of diagnosis), while linear regression was applied to continuous phenotypes (gestational age and blood pressure measurements). All analyses were corrected for batch and the first three principal components were included as continuous covariates. With this analysis we found several strong associations of SNPs to secondary maternal phenotypes (Table V; Supplementary table 12). These findings point to several genetic regions containing relevant genes associated with pregnancy and the complex biology of preeclampsia, as detailed below.

### Gestational Age

Gestational age was associated in mothers with one locus on chromosome 1 (rs952593, beta - 1.66, 95% CI ± 0.61, p=3.12×10^−7^, MAF 0.13). This region is near *TBX15* (Table V; Supplementary Table 12; Supplementary Figure 17-20), a t-box transcription factor shown to be downregulated in intrauterine growth restricted placentas (Chelbi et al., 2011). The association held true with both measurements of gestational age (by maternal last period and neonate Capurro test). The maternal measurement, but not the fetal measurement, of gestational age was associated with a multigenic locus on chromosome 11 (top SNP rs2581927, beta -2.03, 95% CI ± 0.85, p = 4.85×10^−6^; MAF 0.06). A gene of interest in this locus is APLNR, the receptor to ELABELA, which causes preeclampsia symptoms in mice (Supplementary Figures 21-22) (Ho et al., 2017).

### Diastolic and Systolic Blood Pressure

Diastolic blood pressure reached genome-wide significance for one association in the maternal genome on chromosome 4 (top SNP rs1874237, p<5×10-8, beta -4.257, 95% CI -5.711 ─ -2.804, MAF 0.45; Table V; Figure 3). This SNP is within an uncharacterized non-coding RNA locus near *NKX6-1*, a gene involved in β-cell development and function (Taylor et al., 2013). In the offspring, both systolic and diastolic blood pressure were strongly associated with SNPs in *KCNS3/K(V)9.3* (top SNP rs4553827, beta 7.44, 95% CI ± 2.82, p = 5.26×10^−7^, MAF 0.25), a voltage-gated potassium channel gene that is highly expressed in the human placenta, where it localizes to placental vascular tissues and syncytiotrophoblast cells (Fyfe et al., 2012) (Supplementary Table 13; Supplementary Figures 23-26).

**Figure 3.**
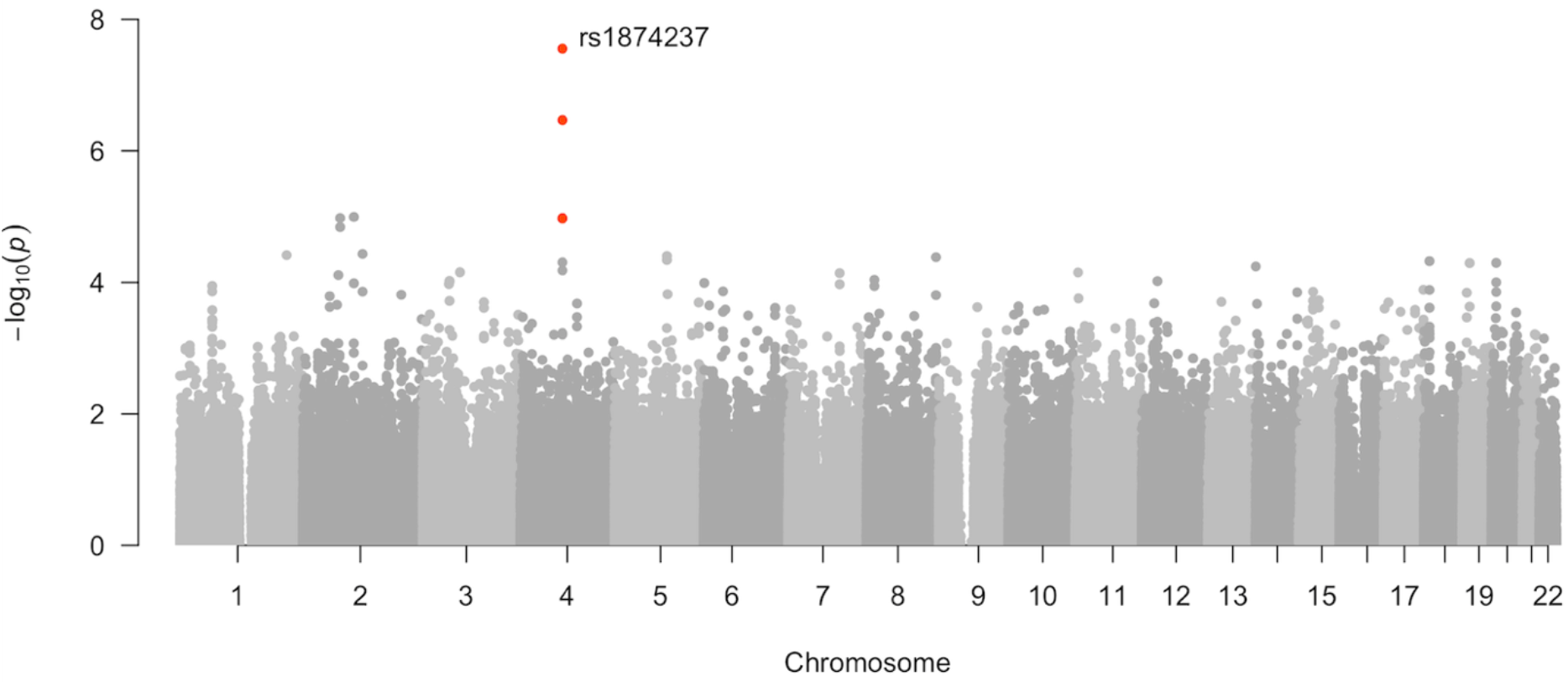
Manhattan plot showing top association in the maternal genome with diastolic blood pressure. SNP rs1874237 on chromosome 4 at p<5×10-8, genomewide significance (shown in red).

### Proteinuria and Severity of Diagnosis

Proteinuria was most strongly associated in the mothers with rs2760751 on chromosome 17 (OR 2.83 ± 1.02, p = 5.65E-06, MAF 0.29). This SNP is intronic to SMG6, a telomerase binding protein. A second association with proteinuria in the maternal genome was found with SNP rs12276362 (OR 0.41 ± 0.14, p = 1.19E-05, MAF 0.49) in chromosome 11, by the *PIWIL4* gene (Supplementary Figures 27-30). This region is also correlated with severity of diagnosis in the mothers (rs1940640, OR 2.4 ± 0.8, p = 1.30E-05, MAF 0.43; Supplementary Figures 29, 31). It is not surprising that proteinuria and severity of diagnosis share a common association, since these two phenotypes are correlated—clinically severe cases generally have higher levels of protein in the urine. Aberrant PIWI proteins, which interact with pi-RNAs to drive post-transcriptional gene regulation, have been found in cancers (Wang et al., 2016), and theoretical evidence from piRNA evolution suggests a role in placentation, although this has yet to be proven empirically (Chirn et al., 2015). In the offspring genome, proteinuria showed an association with placental gene *RARB*, or retinoic acid (RA) receptor beta (rs4241542, OR 0.26 ± 0.14, p=7.04×10^−6^, MAF 0.21) (Comptour et al., 2016, Huebner et al., 2018), while the strongest association with proteinuria is on a different region, in a SNP intronic to STK32B (rs62297274 (OR 0.35 ± 0.12, p=3.5104×10^−6^; Supplementary Table 13; Supplementary Figure 32-33). Interestingly, the minor allele for SNP rs62297274 is found at high frequencies in Peruvians compared to other global populations. In the Puno cohort MAF for this variant is 0.49, slightly higher than among Peruvians from Lima sampled in the 1KG (PEL MAF 0.41) (Supplementary Figure 34). In contrast, the minor allele is found at low frequencies in the rest of the Americas (1KG AMR MAF 0.19) and is rarely observed globally (1KG MAF <0.05) (Supplementary Table 9).

## Discussion

In this analysis, we investigate the genetic diversity of a preeclampsia cohort of Andean families from Puno, Peru; a population with one of the highest incidences of this disease in the world (Bristol, 2009, Gil Cipirán, 2017). We harness the power of a trio study design to uncover maternal, paternal, and fetal genetic factors influencing the incidence and severity of preeclampsia in this cohort. In contrast to previous preeclampsia GWAS studies, which have been hampered by limited phenotyping and heterogeneous sampling (Williams and Broughton Pipkin, 2011), the present work includes a case-control cohort sampled from a single population, treated at the same hospital, and exposed to similar selective pressures due to long-term residence at high altitude. Thus, despite a small sample size, our family based GWAS design permits identification of novel significant and suggestive associations with preeclampsia that would remain otherwise undiscovered (Tishkoff, 2015).

Most genetic studies on preeclampsia have not investigated whole family units (Boyd, et al., 2013, Cincotta and Brennecke, 1998, McGinnis, et al., 2017, Salonen Ros et al., 2000), despite the evidence of a complex genetic risk involving factors from both parents and the fetus (Valenzuela, et al., 2012). This reinforces the strength of our approach, where the top association in the trio study was rs5960, an intronic variant in the clotting factor gene *PROZ*, in a locus with two other clotting factors: *F7* and *F10*. PROZ, a vitamin K-dependent factor, is an anticoagulant protein with a role in factor X inhibition (Almawi et al., 2013). Several previous studies have suggested a hypercoagulative state in preeclampsia (reviewed in Ismail and Higgins, 2011), as spiral arteries of preeclamptic pregnancies often present thrombosis and atherosis (Haram et al., 2014). In fact, strong evidence supporting an effect of thrombotic processes on preeclampsia is based on the observation that aspirin, a known blood thinner, successfully delays preeclampsia onset (Wright and Nicolaides, 2019).

Low PROZ levels are associated with thrombotic disorders, and many adverse pregnancy outcomes have also been linked with maternal PROZ levels (Almawi et al., 2013). A small, prospective case-control study found low PROZ levels associated to intrauterine growth restriction (IUGR) and intrauterine fetal demise, but not preeclampsia (Bretelle et al., 2005). In contrast, a larger cross-sectional study found lower median levels of PROZ in preeclampsia outcomes but not IUGR or fetal demise (Erez et al., 2007). One study found a correlation between lower PROZ levels and severity of HELLP syndrome (a complication of preeclampsia that stands for haemolysis, elevated liver enzymes, and low platelets), which occurs in 10-20% of preeclamptic pregnancies (Haram, et al., 2014, Kaygusuz et al., 2011). However, no study on PROZ or other clotting factors in preeclampsia has been successfully replicated, likely due to the extreme heterogeneity of the disease and the mix of populations studied.

As most previous studies on PROZ have focused on the mother’s genome (Erez, et al., 2007, Xu et al., 2018), ours is the first study to suggest a correlation between the fetal *PROZ*/*F7*/*F10* locus on chromosome 13 and preeclampsia. In a subset of our sample, we found no differences in protein plasma levels of PROZ between preeclamptic and healthy pregnancies in the mother or the offspring. However, this analysis was limited by small sample size and post-natal blood sampling. In other words, since samples were only collected immediately after birth, we were unable to monitor changes in PROZ protein levels throughout the pregnancy. Further longitudinal studies could analyze clotting factor levels and activity in this pregnant population to assess the impact of thrombosis in preeclampsia risk among Andean highlanders.

Expanding the TDT to a parent of origin analysis (POO), we found several associations to genetic regions with suggested paternal inheritance. For instance, the top TDT hit on F10, rs5960, is also the locus with the strongest paternal origin effect in the TDT-POO. Although future research examining variation at the *PROZ/F7/F10* region in a larger population will be needed to confirm this finding, our results are of interest to studies investigating the role of paternal genetic factors, genomic imprinting and paternal-offspring conflict in preeclampsia and other pregnancy disorders (Christians et al., 2017, Galaviz-Hernandez, et al., 2018, Hollegaard et al., 2013, Pilvar et al., 2019, Wikstrom et al., 2012, Zadora et al., 2017).

Other top regions in the TDT-POO include biologically relevant genes SPAG6 and ADGRG6, previously described as being involved in infertility and the immune system (SPAG6) (Cooley, et al., 2016, Neilson, et al., 1999), or angiogenesis (ADGRG6/GPR126) (Cui, et al., 2014, Sitras, et al., 2009). Of these, only ADGRG6 has been associated with preeclampsia in previous research that found it upregulated in preeclamptic placentas (Cui et al., 2014, Sitras et al., 2009). Future work could investigate potential roles of these candidate genes in the maternal-fetal interface and elucidate their involvement in the pathophysiology of preeclampsia.

We also found several placental genes associated with secondary phenotypes that underline the severity of preeclampsia, such as hypertension, gestational age, and proteinuria. Differential expression of these genes may contribute to the insufficiency of placental development in early pregnancy that leads to hypertension and proteinuria in the third trimester. Some of our suggestive associations are near genes previously shown to have roles in pregnancy, vascular processes, and even preeclampsia. One such gene is APLNR, the receptor to ELABELA, which causes preeclampsia symptoms in mice (Ho, et al., 2017) and is lower in the serum and placentas of some women with late-onset, but not early-onset preeclampsia (Zhou et al., 2019). However, this gene is in a multigenic locus, and fine-mapping approaches with functional studies are required to discover the effect of this locus in our cohort.

Our study is one of only a few preeclampsia GWAS studies to include the offspring genome. One recent study with a large cohort found a gene, sFLT1, associated with late (but not early) preeclampsia (Gray, et al., 2018, McGinnis, et al., 2017), suggesting that dysregulation of genes in the fetal genome contribute to preeclampsia. In our study, we found novel fetal associations with preeclampsia and its severity phenotypes in the fetus. For instance, we found an association between severity of hypertension (systolic and diastolic pressure measurements) and KCNS3/K(V)9.3 a gene that is highly expressed in the human placenta, where it localizes to placental vascular tissues and syncytiotrophoblast cells (Fyfe, et al., 2012). We also found an association of the retinoic acid (RA) signaling gene RARB and severity of the proteinuria in the preeclamptic fetal genome. RA signaling is essential for healthy placental and fetal development in animal models, with evidence of similar requirement in humans (reviewed in (Comptour, et al., 2016)). RARB is expressed in the extravillous part of the placenta and its activation induces RARRES, shown in one study to be overexpressed in preeclamptic placentas (Huebner, et al., 2018). Our study adds to this body of literature and highlights the role of RA in proper placentation. Lastly, the most interesting region in the offspring genome was identified in our case-control study; the S100P gene, a calcium-binding protein strongly expressed in the placenta (Zhu, et al., 2015) that promotes trophoblast proliferation in culture (Zhou, et al., 2016). This finding suggests that fetal biology, and specifically placental development driven by fetal genes, highly contributes to the pathology of preeclampsia.

We examined the global distribution of allele frequencies for each of the candidate associated SNPs detailed above. Most alleles were shared among several global populations (see global distribution plots in Supplementary Figures). A notable exception is SNP rs62297274, an intronic variant located in gene STK32B which is associated with proteinuria in the offspring genome. The minor allele reaches its highest global frequency in Peruvian populations (Supplementary Figure 34). As of this writing SNP rs62297274 has no reported clinical significance in dbSNP. However, intronic variants are known to have functional impacts on RNA splicing patterns (Cooper, 2010). To elucidate the functional significance of this variant, future research could evaluate its pathogenic potential in Peruvian populations (Joynt et al., 2020, Lin et al., 2019).

As discussed, several genes found in our analyses are involved in placental function. Interestingly, morphological studies comparing placentas from Andean-descent and European-descent individuals in Bolivia, at both low and high altitudes, describe differences in placental composition (Jackson et al., 1987, Jackson et al., 1988). Highland placentas from individuals of both ancestries show more intervillous space but less villi, and the Andean highland placenta, compared to the European, have more trophoblast and villous stroma on average. Differences in placental morphology suggest an adaptive mechanism to the lower oxygen pressure at high altitude, but one that does not lower the risk of preeclampsia.

In conclusion, this study investigates a cohort of preeclamptic Highland Andean families from Puno, Peru to elucidate the genetic basis of this pregnancy disorder at high altitudes. We generated high-density genotype data at over 400,000 positions across the genome and used these data to determine ancestry patterns and map associations between genetic variants and preeclampsia phenotypes. Our trio-based recruitment strategy, including genotype data from mothers, fathers, and offspring, allowed us to identify novel genetic regions not previously reported in preeclampsia genome-wide association studies. Specifically, we identified strong associations with several variants near genes involved with placental and blood vessel function, and therefore, of functional importance for human pregnancy biology. The strongest association hit involves a cluster of clotting factor genes on chromosome 13 including *PROZ*, *F7* and *10* in the fetal genome. This finding provides supporting evidence that coagulation plays an important role in the pathology of preeclampsia and potentially underlies other pregnancy disorders exacerbated at high altitude.

Studying diverse human groups with unique genetic adaptations enables identification of the primary genetic factors underlying complex phenotypes and gene function. This research examined Andean populations as a model to understand human pregnancy physiology in hypoxic conditions. This natural experimental setting provides a unique opportunity to understand the genetic factors influencing human reproductive fitness in challenging environments worldwide and to discover population-specific variants underlying biomedical traits. Our work also underscores the importance of including diverse populations in genome wide association studies and functional variant discovery efforts to better understand human physiology and disease globally.

## Data Availability

The data underlying this article are available in the European Genome-Phenome Archive (EGA) at https://ega-archive.org/ and can be accessed with Data Access Committee approval under Study EGAS00001004625.

## Authors’ roles

K.M.B.R. and M.A.N.C. wrote the article with input from G.L.W., A.M.E. and J.C.B. K.M.B.R., M.A.N.C., J.W.C., E.T.Z., C.R.G. and G.L.W. performed data analyses. P.O.T. and K.S.M. designed and coordinated data collection. P.O.T., K.S.M, L.E.L., V.V.D., J.C.M.C., F.M.C. and G.P.Y.P. collected samples and medical records in Puno and handled fieldwork logistics. K.M.B.R., M.A.N.C., A.S., E.R., G.M.H., R.C.S, R.C., C.E., S.H., E.G.B., E.T.Z., G.P. and C.G. performed laboratory work. C.D.B, J.C.B., C.R.G., A.M.E., C.G., and M.A.N.C. provided resources, funding, and/or laboratory space. All authors revised the article and approved the final submitted version.

## Acknowledgements

We extend our deepest gratitude to the people of Puno, Peru who participated in this study at *Hospital Regional Manuel Nuñez Butrón* and *Universidad del Altiplano*. We are tremendously grateful to Javier Mendoza Revilla who provided commentary on the final version of this manuscript, and to the Mendoza Revilla family, who provided lodging and logistics support in Lima during fieldwork seasons of the technical team.

## Author notes

Keyla M. Badillo Rivera and Maria A. Nieves-Colón contributed equally as first authors. Christopher R. Gignoux, Genevieve L. Wojcik and Andrés Moreno-Estrada contributed equally as last authors.

